# Developing a Stakeholder-Driven One Health Collaboration Framework for Bovine Tuberculosis Control at the Human-Animal Interface in Lundazi and Monze Districts, Zambia

**DOI:** 10.64898/2026.11.23.26368721

**Authors:** Anthony Phiri, Mildred Zulu, Maxwell Phiri, Wezi Nyirenda, Enos Phiri, Francisco Olea-Popelka, Sydney Malama

## Abstract

**Background:** Bovine tuberculosis, caused by Mycobacterium bovis, is a neglected zoonosis at the human-animal interface in Zambia, particularly affecting cattle-dependent rural communities in Eastern and Southern Provinces, where cattle are central to livelihoods, nutrition, and cultural identity. While Zambia has a One Health Strategic Plan 2022-2026 and a One Health Secretariat under Zambia National Public Health Institute, operational collaboration between veterinary and medical sectors at district level remains fragmented, contributing to diagnostic gaps, underreporting, and continued transmission through consumption of raw milk, undercooked meat, and occupational exposure during slaughter and handling of infected carcasses. The disconnect between national policy and district implementation has created a vacuum where frontline workers operate in isolation despite sharing the same disease burden.

**Methods:** This qualitative exploratory study was conducted in Lundazi District (Eastern Province) and Monze District (Southern Province), Zambia, from February to June 2022. Ten Focus Group Discussions were conducted with 104 participants, including cattle farmers, cattle businessmen, abattoir workers, veterinary and para-veterinary professionals, and medical doctors, and ten Key Informant Interviews were conducted with District Veterinary Officers, District Medical Officers, Ministry of Health officials, and Ministry of Fisheries and Livestock officials. Participants were purposively selected based on their role, experience, and knowledge of bovine tuberculosis and One Health collaboration. Discussions were held in local languages (Tumbuka, Tonga, Chewa), audio-recorded with consent, transcribed verbatim, translated into English, and imported into NVivo 12 (QSR International, Melbourne, Australia) for thematic analysis following Braun and Clarke’s six-step approach, with framework matrices for triangulation and trustworthiness. This methodological rigor ensured that diverse voices from community to policy level were systematically captured and compared.

**Results:** Five themes emerged: (1) awareness of bovine tuberculosis, (2) socio-economic impacts, (3) public health impacts, (4) cultural practices and transmission, and (5) collaboration gaps and proposed framework. Medical and veterinary professionals demonstrated excellent awareness, while cattle farmers and abattoir workers showed poor awareness. Socio-economic impacts included reduced milk yield and carcass condemnation. Key barriers to collaboration included lack of diagnostic capacity at district hospitals to differentiate Mycobacterium bovis from Mycobacterium tuberculosis, siloed training curricula, absence of formal round-table coordination mechanisms, and limited funding. An existing enabler was informal collaboration in meat inspection between veterinary assistants and environmental health officers. Based on these findings, a five-pillar One Health Collaboration Framework is proposed. This framework was co-constructed from participant recommendations, making it contextually grounded and acceptable to end-users.

**Conclusion:** Effective control of bovine tuberculosis in Zambia requires institutionalized One Health collaboration, moving from informal ad hoc interactions to formal governance structures. The proposed framework provides a practical, low-cost and contextually grounded roadmap for district-level implementation and is applicable to other zoonoses such as brucellosis, anthrax and rabies. Its implementation can help break the cycle of under-reporting and under-funding that currently sustains zoonotic transmission in rural Zambia.

## INTRODUCTION

Bovine tuberculosis (bTB) is a chronic granulomatous disease caused by Mycobacterium bovis, a [1] member of the Mycobacterium tuberculosis complex, affecting cattle, wildlife and humans. Globally, zoonotic tuberculosis accounts for an estimated 140,000 new human cases and 12,000 deaths annually,[2] with Africa bearing a disproportionate burden due to close human-animal interactions, consumption of unpasteurized dairy products, and limited diagnostic capacity to differentiate zoonotic from human tuberculosis. Transmission occurs through ingestion of contaminated milk and dairy products, consumption of undercooked meat containing tuberculous lesions, inhalation of infectious aerosols generated during slaughter, and direct contact through cuts and abrasions during handling of infected carcasses and offal.[1][2] These transmission pathways are amplified in settings where personal protective equipment is rarely used and where slaughter is conducted in open, poorly ventilated spaces. [3]

In Zambia, cattle farming is central to rural livelihoods, food and nutrition security and cultural practices in Eastern and Southern Provinces. The Livestock Annual Census reported 63,144 cattle in Lundazi and 284,713 in Monze districts, contributing to provincial herds of 597,149 and 1,225,090 for Eastern and Southern Provinces, respectively. Cattle provide milk, meat, draft power, manure and serve as a store of wealth for paying school fees and health services. In these settings, human-animal interaction is intimate and daily, creating a high-risk interface for zoonotic transmission[4]. Previous studies in Zambia have documented high-risk practices including consumption of unpasteurized milk as a sign of hospitality, consumption of undercooked meat and handling beef with bare hands, and economic losses due to reduced milk yield and carcass condemnation at abattoirs. Such losses disproportionately affect women and children who depend on milk for household nutrition, thereby linking animal disease directly to human nutritional vulnerability.[5]

The One Health approach is defined as an integrated, unifying approach that aims to sustainably balance and optimize the health of people, animals and ecosystems, recognizing that health of humans, domestic and wild animals, plants and wider environment are closely linked and interdependent.[6] It is advocated by the Quadripartite composed of Food and Agriculture Organization, United Nations Environment Programme, World Health Organization and World Organisation for Animal Health as essential for controlling zoonoses at the human-animal-environment interface.[7][8] The Quadripartite has developed the One Health Joint Plan of Action 2022-2026, and the Tripartite Guide to Addressing Zoonotic Diseases provides practical steps for operationalization. Zambia launched its One Health Strategic Plan 2022-2026 and established a One Health Secretariat under Zambia National Public Health Institute to coordinate multisectoral collaboration, yet district-level operationalization remains weak due to lack of formal structures, separate budgets, siloed training curricula, and the absence of joint platforms. [8] [9] [10] Similar challenges of siloed surveillance, separate funding streams, and lack of joint outbreak response have been documented in Tanzania,[10] where One Health platforms exist nationally but not sub-nationally, in Ethiopia[11] where a systematic review showed diagnostic gaps, and in Uganda where separate ministries have separate goals and resource competition hinders collaboration. Without addressing these operational bottlenecks, national strategies risk remaining aspirational documents rather than functional tools for disease control.[12]

While quantitative data can quantify burden and awareness gaps, qualitative inquiry is needed to understand why collaboration fails, what cultural beliefs and socio-economic factors drive transmission, what existing informal collaborations exist, and how to design a contextually appropriate, low-cost framework that is acceptable to frontline implementers. To date, no study in Zambia has developed a stakeholder-driven One Health collaboration framework specifically for bovine tuberculosis control using qualitative methods at district level involving both community members and policy actors. This study addresses this critical gap. The objective of this study was to explore barriers and enablers to collaboration between veterinary and medical sectors and to develop a stakeholder-driven One Health collaboration framework for controlling bovine tuberculosis and other zoonoses in Zambia.[13]

## METHOD

### Study design

This qualitative exploratory study was conducted in Lundazi District (12.2849°S, 33.1745°E), Eastern Province, and Monze District (16.2803°S, 27.4733°E)[14], Southern Province, Zambia, from February to June 2022. Lundazi comprises nine study villages: Engeleweni, Kacindila, Efumbeni, Gowa, Themba, Katoto, Chizungu, Chipetuka and Bokosi. Monze comprises eleven villages: Munababa, Mazunkha, Shimbayi, Mwanakaba, Hamusonde, Shimbwaalale, Mwando, Chavwa, Hankogote, Hashote and Chilomo. Lundazi District borders Kasungu National Park in

Malawi, while Monze District lies northwest of Lochinvar National Park with a transhumance pastoral system into Kafue flats where cattle interact with Kafue lechwe antelope (Kobus leche kafuensis), a known wildlife reservoir for M. bovis, creating wildlife-livestock-human interfaces known as hotspots for bovine tuberculosis transmission. The two districts were selected due to their traditional livestock systems, high cattle populations, and proximity to wildlife interfaces. Their selection also reflects contrasting agro-ecological zones, allowing comparative insights into different transmission ecologies.[14][15]

### Participant selection

Participants were purposively selected based on their role, experience, knowledge, and involvement in bovine tuberculosis control and One Health collaboration [16]. The aim was to include diverse perspectives across the human-animal interface, from community to policy level, ensuring that both technical and lived experiences were represented.

#### For Focus Group Discussions, 104 participants were included

Cattle farmers who are primary cattle keepers, cattle businessmen who trade cattle at markets recruited via snowball sampling at cattle markets, abattoir workers including slaughtermen and meat inspectors, veterinarians and para-veterinary professionals including veterinary assistants and environmental health officers, and medical doctors and clinicians working in district hospitals.[16] Ten Focus Group Discussions were conducted, five per district, each comprising 8-12 participants and lasting up to two hours[16]. Discussions were stratified by age (18-30, 30-41, 41-50, >50 years) and gender to facilitate open dialogue[16] and ensure participants could speak freely without hierarchy, following established qualitative methodology. This stratification minimized power imbalances that often silence women and younger workers in mixed groups.

For Key Informant Interviews, ten key informants were purposively selected to cover district and national policy levels and to provide triangulation[16]

District Veterinary Officer, Lundazi District (1), District Veterinary Officer Monze District (1), District Medical Officer Lundazi District (1), District Medical Officer Monze District (1), three officials from Ministry of Health headquarters, and three officials from Ministry of Fisheries and Livestock headquarters. Interviews lasted 45-60 minutes[16] and were conducted in English at the participants’ offices. The sample size was determined by the principle of data saturation, where recruitment and analysis continued iteratively until no new themes or insights emerged, with saturation achieved after eight Focus Group Discussions and eight Key Informant Interviews, consistent with Guest and colleagues who found that 90% of themes emerge within six interviews and saturation typically within twelve interviews. Saturation was assessed daily through debriefing meetings among the research team.[16][17][18]

### Data collection instruments

Two semi-structured interview guides were developed based on study objectives, One Health literature, and the Zambia One Health Strategic Plan[19]. One guide focused on One Health collaboration at district and community levels, exploring awareness of bovine tuberculosis, perceived socio-economic and public health impacts, cultural practices influencing transmission, and experiences of collaboration between veterinary and medical sectors[19]. The second guide focused on policy implementation for national-level officials, exploring governance, funding, and operationalization of One Health strategies[19]. Guides included open-ended questions and probes. Guides were translated into local languages (Chewa, Tonga, Tumbuka) by language experts and back-translated to English to ensure consistency and cultural appropriateness[19]. All Focus Group Discussions and Key Informant Interviews were conducted by the principal investigator, who is fluent in English and familiar with local languages, supported by a local-language translator, audio-recorded with written consent, and field notes were taken on non-verbal cues and context[19]. Document review of bovine tuberculosis regulations, meat inspection guidelines, and One Health policy documents was conducted to corroborate primary data and enhance credibility. The review also included district health reports, laboratory registers and abattoir condemnation records[19].

Pilot testing of the guides was conducted in Chisamba District, Central Province, with participants similar to the study population to assess clarity, cultural appropriateness, flow, and duration, and guides were revised accordingly. No invasive procedures were performed.[19]

### Data analysis

All audio files from Focus Group Discussions were translated into English where necessary and transcribed verbatim by the research team. Transcripts were read repeatedly to achieve immersion and familiarization. Data were imported into NVivo 12 (QSR International Pty Ltd, Melbourne, Australia) for data management and coding[20]. Thematic analysis followed Braun and Clarke’s six-phase approach: (1) familiarization with data, (2) generating initial codes, (3) searching for themes, (4) reviewing themes, (5) defining and naming themes, and (6) producing the report. Initially, broad coding was performed using major themes derived from the topic guide, followed by a second round of inductive coding into sub-themes emerging from data. Framework matrices were constructed, extracting key quotations under each sub-theme and summarizing narratives for each participant group. These matrices were used to cross-check narratives from Key Informant Interviews with Focus Group Discussion participants to identify convergent or divergent views and enhance triangulation[20]. Illustrative quotations representing themes are presented with participant identifiers to preserve anonymity. To ensure trustworthiness, credibility was enhanced through prolonged engagement, triangulation of data sources and member checking; dependability through an audit trail; and confirmability through reflexive journaling[18][21]. Reporting followed the Consolidated Criteria for Reporting Qualitative Research (COREQ) 32-item checklist for interviews and focus groups[18].Coding was independently verified by two researchers to improve inter-coder reliability. [20][21]

### Ethics statement

Ethical approval was obtained from the University of Zambia Biomedical Research Ethics Committee (UNZABREC Ref: 2102-2021). Permission was obtained from Provincial and District Veterinary and Health Offices and local chiefs. Written informed consent, including thumbprint for participants unable to write, was obtained after explaining the study purpose, voluntary nature, confidentiality, anonymity, and right to withdraw at any time without penalty. No compensation was provided. Identifiers were replaced with numeric codes, and data were stored in encrypted, password-protected files and locked cabinets accessible only to the research team

## RESULTS

Five themes emerged from the analysis: (1) awareness of bovine tuberculosis, (2) socio-economic impacts, (3) public health impacts, (4) cultural practices and transmission, and (5) collaboration gaps and proposed framework. This manuscript focuses on Theme 5, with Themes 1-4 summarized for context.

**Table 1:** FGDs = Focus Group Discussions (10 groups, 104 participants: 42 Lundazi, 62 Monze). KIIs = Key Informant Interviews. DMO = District Medical Officer. DVO = District Veterinary Officer. EHT = Environmental Health Technologist. MoU = Memorandum of Understanding. DHIS2 = District Health Information System 2. bTB = bovine tuberculosis. Source Authors field data

| **Theme** | **Community Perspective: FGDs (n=104 farmers, butchers, and abattoir workers)** | **KIIs (n=10, District & National officials)** | **Document Evidence & Convergence Assessment** |
|---|---|---|---|
| Diagnostic capacity gap | "We see lesions but no lab. We throw away" - Abattoir workers FGD Lundazi. "Milk is milk, we drink" - Farmers Monze | "We do not have instruments to test bovine TB... And because we report zero cases, we get no funding - so no instruments. It is a cycle" - DMO Monze, June 2022 | Evidence: District lab registers 2022: 0 bTB tests done. DHIS2: 0 zoonotic TB cases reported from 2 districts. Lab inventory: No GeneXpert cartridges for bTB, no culture media. Convergence: STRONG CONVERGENCE - All three sources confirm diagnostic gap. Explains persistence of zoonotic risk. |
| Absence of joint One Health platform | "Never seen them together - doctor and vet doctor - they never come together" - Farmers FGD Lundazi. "Each one works alone" - Farmers Monze | "No round table for medical and veterinary... this is alarming because disease doesn't know boundary" - DVO Monze, Feb 2022. "We plan separately" - DMO Lundazi | Evidence: Reviewed district files: No Memorandum of Understanding between MOH and MFL at district, no joint workplan, no joint minutes 2021-2022, no joint budget line. National One Health Strategic Plan 2022-2026 found at provincial office but not disseminated to district. Convergence: IMPLEMENTATION GAP - Policy exists at national level but not operationalized at district level. |
| Enabler: Existing informal collaboration at meat inspection | "Vet assistants come every morning to check our meat before selling" - Butchery owners Monze FGD | "Meat inspection is done between vet and environmental - we collaborate there" - DVO Lundazi & EHT Monze KII | Evidence: Public Health Act, Food Safety Act, Meat Inspection Guidelines mandate joint inspection. Abattoir registers show joint signatures of Vet Assistant and EHT. CONVERGENCE: Strong convergence on enabler - This is the only existing One Health practice at the district and can be leveraged as an entry point for integrated bTB surveillance and community education. |

Themes 1-4 summary: Medical and veterinary professionals demonstrated excellent awareness, while cattle farmers and abattoir workers showed poor awareness; socio-economic impacts included reduced milk yield and carcass condemnation, and cultural practices included consuming raw milk as hospitality and handling meat without gloves, increasing zoonotic risk. Full details of Themes 1-4 are reported separately, with Theme 5 the focus of this manuscript.

### Theme 5: Collaboration gaps and proposed framework

#### Diagnostic capacity gap

All District Medical Officers and District Veterinary Officers reported no capacity to diagnose zoonotic tuberculosis at district hospital and veterinary laboratory levels, leading to under-reporting and a cycle of limited financial support and low prioritization. Participants described that tissue samples must be sent to central laboratories in Lusaka, causing delays and loss to follow-up. This theme was consistent across both districts and across both human and animal health sectors. The absence of molecular tools such as GeneXpert with M. bovis differentiation capacity at district level further compounds the problem, leaving clinicians to treat empirically without confirming zoonotic origin.

“*We do not have the instruments and the capacity to diagnose Bovine Tuberculosis and other zoonotic diseases. Worse still, if we cannot produce the number of cases of Bovine Tuberculosis and other zoonotic diseases, then there will be little financial support. Alternatively, if we were able to give such statistics, it would prove to people that they should have an interest in such diseases and therefore put funding towards diagnostic infrastructure*”

[District Medical Officer, Monze District, June 2022].

*“I strongly don’t believe that the laboratory at the district hospital has the full capacity to diagnose Bovine Tuberculosis from tissue samples, and instead it would need to be sent to Lusaka*”

[District Medical Officer, Lundazi District, February 2022].

*“Many times we learned about the way to diagnose Bovine Tuberculosis at some point in our training, but since we do not see it often, our ability to diagnose it is minimal”*

[District Medical Officer, Monze District, February 2022].

*“There is very little funding allocated to Bovine Tuberculosis and other zoonotic diseases, with limited funding, it becomes a challenge to distinguish Bovine Tuberculosis from other diseases because they are carrying similar clinical signs. Bovine Tuberculosis is similar to fever and brucellosis*

[District Medical Officer, Lundazi District, June 2022].

#### Siloed training and absence of joint platforms

Participants described separate training curricula for medical and veterinary professionals, separate reporting systems, separate budgets, and the absence of joint planning forums or round-table meetings to brainstorm and execute control measures, which was described as alarming. Resource competition was highlighted as a barrier. This siloed approach means that even when both sectors recognize the same outbreak, they respond in parallel rather than jointly, wasting scarce resources and missing opportunities for integrated surveillance

*“The veterinarians and the medical officers have been trained in different areas of tackling diseases. However, there has not been any round table meeting to brainstorm and to execute the ideas gained during the workshops to fight the disease, which is alarming in the district*

[District Veterinary Officer, Monze District, February 2022].

*“The major challenge is the issue of resources. Many times, when medical personnel have resources, they are only concerned with diseases that are not zoonotic. But here, since we are in animal and public health, we want to collaborate. Though collaboration hasn’t happened much since both sources don’t have the same goals”*.

[District Veterinary Officer, Lundazi District, 2022].

#### Existing informal collaboration – Enabler

Despite gaps, an existing informal collaboration was identified as an enabler and potential foundation for formalization, where meat inspection is conducted jointly by veterinary assistants and environmental health personnel. This daily interaction at abattoirs and slaughter slabs represents a natural One Health touchpoint that is already trusted by communities and does not require additional infrastructure.

*“There seems to be some collaboration where meat inspection is done between veterinary assistants and environmental personnel. This applies in diagnosing zoonotic Tuberculosis, the medical personnel are involved”*

[District Veterinary Officer, Lundazi District, 2022].

#### Proposed Five-Pillar One Health Collaboration Framework (Fig 1)

Based on qualitative findings and participant recommendations, a framework for collaboration between veterinary and medical experts is proposed (Fig 1). The framework is intentionally low-cost and leverages existing personnel rather than creating parallel structures.

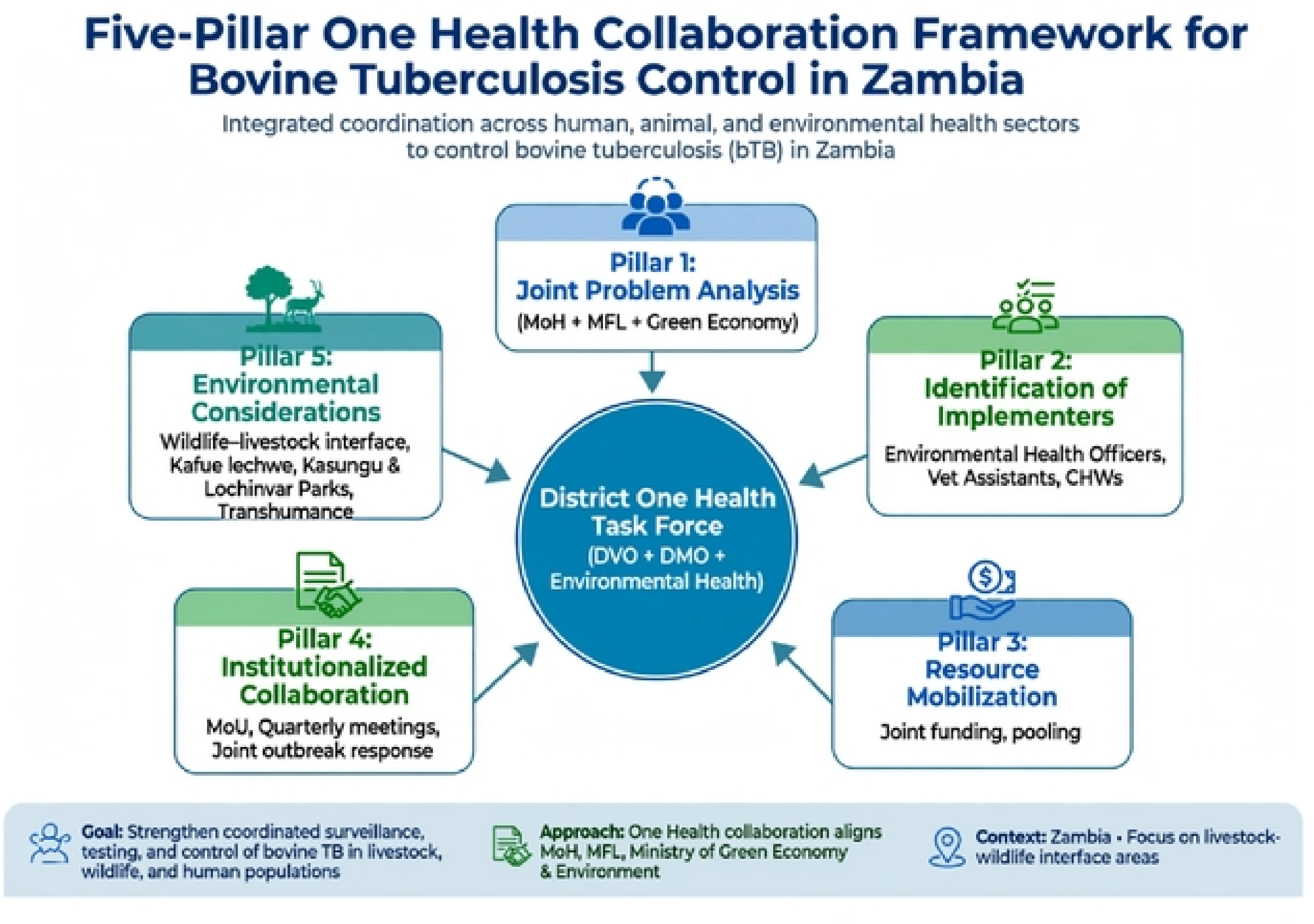
Five-Pillar One Health Framework

Pillar1: Joint Problem Analysis – identify and prioritize problems together between Ministry of Health, Ministry of Fisheries and Livestock and Ministry of Green Economy and Environment through joint meetings. This involves quarterly joint risk mapping and participatory disease surveillance involving community leaders.

Pillar 2: identify and clarify roles of implementers on the ground including environmental health officers, veterinary assistants and community health workers to simplify monitoring and evaluation. Clear terms of reference and joint accountability mechanisms are proposed.

Pillar 3: Resource Mobilization—identify reliable sources of funding for environmental conservation and implementation, including joint proposal writing and pooling resources. This includes integrating bTB activities into existing District Health and Livestock annual budgets and leveraging donor alignment through the One Health Secretariat.

Pillar 4: Institutionalized Collaboration—establish a multidisciplinary District One Health Task Force comprising veterinary experts, medical professionals, and environmental health officers with a formal memorandum of understanding, quarterly meetings, and joint outbreak response teams to coordinate disease control efforts and promote One Health.

Pillar 5: Environmental Considerations—integrate environmental factors into disease surveillance, diagnosis, and control measures, recognizing the impact of environmental changes, wildlife-livestock interactions, and transhumance on human and animal health. This pillar explicitly links climate variability, grazing patterns, and wildlife movements to disease risk.

## DISCUSSION

This qualitative study provides the first stakeholder-driven One Health collaboration framework for bovine tuberculosis control in Zambia, grounded in perspectives from frontline implementers and policymakers in Lundazi and Monze districts. Our findings reveal a diagnostic capacity paradox: while professional awareness is excellent among doctors and veterinarians, diagnostic capacity at district level is absent, leading to a vicious cycle of no data, no funding, and no capacity, as articulated by the District Medical Officer in Monze. This under-reporting leads to low prioritization in district health planning and perpetuates neglect. This mirrors findings from Ethiopia, where a systematic review identified a lack of laboratory capacity to differentiate Mycobacterium bovis from Mycobacterium tuberculosis as a key gap[10], and from Tanzania, where district laboratories lack tools for zoonotic tuberculosis diagnosis[9]. The paradox is particularly concerning because it creates a self-reinforcing loop that renders zoonotic TB invisible to decision-makers.[1]

The absence of formal round-table mechanisms and joint platforms explains why Zambia’s One Health Strategic Plan 2022-2026 remains unoperationalized at district level, despite the existence of a national One Health Secretariat[11]. Participants described training in silos, separate institutional mandates, and resource competition as barriers, similar to findings from Uganda, where separate ministries have separate goals and performance indicators, and from Tanzania, where One Health platforms exist nationally but not sub-nationally [11][9]. Our framework’s emphasis on a formal memorandum of understanding between the District Veterinary Officer and District Medical Officer, establishment of a District One Health Task Force including environmental health officers, and joint problem analysis addresses this governance gap, aligning with recommendations for institutionalized collaboration in the Tripartite Guide[6]. Institutionalization is paramount because informal goodwill collapses when staff are transferred, as is common in district systems.[11]

Our five-pillar framework operationalizes the WHO-FAO-WOAH Tripartite Guide to Addressing Zoonotic Diseases and the Quadripartite Joint Plan of Action 2022-2026, but adds contextual specificity critical for Zambia, the transhumance pastoral system in Monze, where cattle seasonally move into Kafue flats, contacting Kafue lechwe, a known wildlife reservoir for M. bovis, proximity to wildlife corridors Blue Lagoon National Park, Lochinvar National Park and inclusion of environmental health officers as core implementers, often overlooked in One Health discussions yet very imperative for meat inspection.[23]

Cultural practices identified, including consumption of unpasteurized milk as a sign of hospitality and respect during traditional ceremonies, are consistent with studies in Southern Province, Zambia. [4] Belief that undercooked meat provides strength, handling meat without gloves because meat is perceived as clean, and belief that BCG vaccination protects against bovine tuberculosis when handling meat with bare hands are consistent with studies in East Africa. [5]

Strengths of this study include use of the Consolidated Criteria for Reporting Qualitative Research (COREQ) 32-item checklist for reporting qualitative research, purposive sampling covering both policy and implementation levels and both human and animal health sectors, triangulation of Focus Group Discussions and Key Informant Interviews and document review, prolonged engagement in study sites, audit trail, and achievement of data saturation, enhancing credibility, dependability, and confirmability. The inclusion of both Tonga, Chewa, and Tumbuka-speaking communities also improves transferability across Zambia’s diverse cultural settings. [20][21][18]

## LIMITATIONS

Limitations include that the study covered only two districts in Eastern and Southern Provinces, so findings may not be generalizable to all settings across Zambia with different livestock systems but are transferable to similar agro-pastoral settings with wildlife-livestock-human interfaces. The study focused on bovine tuberculosis and may not capture dynamics of other zoonoses, though participants identified brucellosis, anthrax, and rabies as priority zoonoses. Social desirability bias may have occurred, as participants may have reported what they thought was expected. To mitigate this, interviews were conducted in neutral venues, and confidentiality was repeatedly emphasized.

For policy, there is a need to formalize a District One Health Task Force with quarterly meetings, a formal memorandum of understanding between Ministry of Health and Ministry of Fisheries and Livestock, joint planning and budgeting, and inclusion of environmental health officers, as recommended by the Zambia One Health Strategic Plan 2022-2026. For practice, there is a need to strengthen district laboratories for zoonotic tuberculosis differentiation using molecular tools, integrate abattoir condemnation data into the human health information system DHIS2, conduct joint outbreak investigations, and develop culturally sensitive joint sensitization materials. For research, there is a need to validate the proposed framework using a Delphi method with 10-15 national One Health experts, assess its cost-effectiveness and feasibility, and expand to other zoonoses such as brucellosis, anthrax, and rabies, which were identified as priority zoonoses by participants. Cost-effectiveness analysis will be essential to convince Treasury to allocate joint budget lines for One Health activities.

## CONCLUSION

Controlling bovine tuberculosis in Zambia requires moving from siloed sectoral approaches to synergistic One Health collaboration. Our stakeholder-driven five-pillar framework, grounded in frontline experts’ perspectives from Lundazi and Monze districts, provides a practical, low-cost, and contextually appropriate roadmap for district-level implementation, addressing diagnostic, governance, funding, and cultural barriers, and building on evidence that cooperation between human and animal health can strengthen health systems. This framework is applicable to other zoonoses and can contribute to improved public health and socio-economic well-being of cattle-dependent communities and achievement of Sustainable Development Goals. [24]

## Data Availability

All relevant data underlying the findings of this qualitative study are within the manuscript and its Supporting Information files. This includes Table 1 triangulation matrix with community perspectives, key informant quotes, and document evidence convergence assessment, the framework description, and participant summary. This study is the qualitative component of a larger mixed-methods thesis the quantitative dataset (n=280 survey) is reported separately and is not part of this manuscript's findings.

## ACKNOWLEDGMENTS

We thank the District Veterinary Officers and District Medical Officers in Lundazi and Monze, chiefs, community health workers, research assistants, and all participants. We thank the University of Zambia Biomedical Research Ethics Committee for ethical clearance.

